# Adenovirus as a Causative Agent of an Acute Gastroenteritis Outbreak in Salvador, Bahia, Brazil

**DOI:** 10.64898/2026.10.19.26365929

**Authors:** Beatriz Peres Vidal, Silvana Oliveira Neves, Ively Paixão Santos, Silvia Ines Sardi, Gubio Soares Campos

## Abstract

**Introduction:** Acute gastroenteritis (AGE) is a cause of childhood morbidity and may be associated with human adenovirus (HAdV) infection. This study aimed to investigate the occurrence of HAdV in fecal samples from children with AGE in Salvador, Bahia, Brazil, as well as to molecularly characterize the identified strains and evaluate their cytopathic effects in cell culture.

**Methods:** A total of 131 samples were analyzed using the nested polymerase chain reaction (Nested-PCR) technique. Molecular characterization was performed through Sanger sequencing and phylogenetic analysis, while Vero cell cultures were used to investigate the occurrence of cytopathic effects.

**Results:** Of the samples analyzed, 13 (9.9%) tested positive for HAdV, while 118 (90.1%) were negative. The highest detection frequency was observed in children aged three years or younger, particularly those under one year of age. Phylogenetic analysis identified variants related to HAdV-F41 and HAdV-B3, revealing genetic differences and distinct clustering patterns compared with reference sequences. In cell culture, cytopathic effects were observed four days after inoculation, although further analyses are required to confirm productive viral replication.

**Conclusion:** These findings demonstrate the presence of HAdV in the samples studied and contribute to understanding its genetic diversity and circulation in Salvador, highlighting the importance of epidemiological surveillance and further studies on its role in the etiology of childhood AGE.

## Introduction

Acute gastroenteritis (AGE) is an inflammatory condition of the intestine caused by pathogens such as adenoviruses. It produces symptoms including diarrhea and vomiting, which may lead to dehydration, particularly in children and older adults (Pessôa Germano Mendes; Vasconcelos Rocha, 2023). Transmission can occur through the consumption of contaminated food or water, as well as through direct contact with individuals infected by pathogens that cause AGE (Cheng et al., 2008; Oba et al., 2020; Pessôa Germano Mendes; Vasconcelos Rocha, 2023).

Acute diarrhea in children is a major cause of morbidity and mortality among those younger than five years and is frequently associated with viral infections (Sociedade Brasileira de Pediatria, 2017). According to data from the Brazilian Ministry of Health, nearly 80,000 diarrhea-related hospitalizations among children were recorded in Brazil in 2018 (SES, 2023). In addition, 558,737 cases of diarrhea were recorded among children younger than ten years between 2000 and 2011, with an overall hospitalization rate of 4.3 per 1,000 children. The rate was even higher among infants younger than one year, reaching 11.2 hospitalizations per 1,000 children (Meneguessi et al., 2015).

Although mortality from acute diarrhea among children younger than five years has declined, diarrhea remains a major cause of childhood morbidity and mortality in Brazil (Meneguessi et al., 2015). These findings underscore the need for interventions aimed at preventing and treating the disease.

Strategies to prevent acute childhood diarrhea include rotavirus vaccination, exclusive breastfeeding during the first six months of life, and appropriate food hygiene practices (Brandt; de Castro Antunes; da Silva, 2015). Understanding the causes of acute diarrhea and its associated risk factors is also essential for improving child health and reducing disease-related mortality.

Despite reductions in mortality associated with gastroenteritis, the disease continues to cause deaths worldwide. In 2019, it was reported as the eighth leading cause of death, accounting for 2.7% of deaths globally (Global Health Estimates, 2021), and it is currently reported as the seventh leading cause in developing countries (Global Health Estimates, 2024). In Brazil, 3,115 deaths attributed to AGE were reported between 2012 and 2020, with the Northeast being the most affected region (Veras et al., 2022). Among Brazilian children, it was reported as the second leading cause of death between 1990 and 2015 (França et al., 2017). These data highlight the need for effective prevention and control strategies to reduce the clinical consequences of the disease.

Human adenovirus (HAdV) was first isolated and characterized in the United States in 1953 in association with respiratory infections (Hilleman; Werner, 1954). The virus belongs to the family Adenoviridae and the genus Mastadenovirus. HAdVs are described as comprising 57 serotypes classified into seven species (A–G). Species F and G are generally associated with acute gastroenteritis in humans, regardless of age (Radke; Cook, 2018; Walker et al., 2013).

Adenoviruses are non-enveloped viruses, approximately 70–90 nm in diameter, with icosahedral symmetry. Their genomes consist of linear, nonsegmented double-stranded DNA (Gallardo et al., 2021), approximately 35–36 kb in length, encoding around 40 proteins. The structural proteins include II (hexon), III (penton base), IIIa, IV (fiber), IVa2, V, VI, VII, VIII, IX, and µ, whereas E1A, E1B, E2, E3, and E4 are nonstructural proteins. Protein V is described as specific to HAdV (Davison; Benkő; Harrach, 2003).

## Materials and Methods

This descriptive cross-sectional study was approved by the Research Ethics Committee of the Institute of Health Sciences, Federal University of Bahia (ICS-UFBA), under CAAE registration number 61239422.9.0000.5662. Viral DNA was analyzed by polymerase chain reaction (PCR). The cycling conditions consisted of an initial denaturation step at 94 °C for 5 min; 30 cycles of 94 °C for 1 min, 54 °C for 45 s, and 72

°C for 2 min; and a final extension step at 72 °C for 5 min (Xu; McDonough; Erdman, 2000). The reactions were performed using GoTaq® Green Master Mix (Promega, USA). The primers used in this study were previously described by Xu et al. (2000).

HAdV PCR amplicons were subjected to electrophoresis on 2% agarose gels prepared in TBE (Tris–borate–EDTA) buffer. After electrophoresis, the gels were stained with ethidium bromide (10 µg/mL) for 5 min and visualized using an ultraviolet transilluminator. The amplified HAdV product was reported to be 482 bp in length.

For HAdV isolation, Vero cells (CCL-81) obtained from the American Type Culture Collection (ATCC) were used. Cells were maintained in Dulbecco’s Modified Eagle Medium (DMEM) supplemented with 10% fetal bovine serum, penicillin, streptomycin, and amphotericin B, at 37 °C in a humidified atmosphere containing 5% CO_2_.

The viral inoculum was prepared by homogenizing 1 g of fecal sample in 5 mL of phosphate-buffered saline (PBS) supplemented with antimicrobial agents, followed by centrifugation at 7,700 rpm for 20 min at 4 °C. The resulting supernatant was used to inoculate Vero cells. For viral isolation, 500 µL of inoculum was added to the cells using a co-infection procedure, followed by incubation at 37 °C for 1 h to allow viral adsorption. After inoculation, cultures were maintained in DMEM supplemented with 2% fetal bovine serum and monitored daily for up to seven days to assess the development of cytopathic effects. Viral infection was subsequently assessed by molecular testing.

For molecular characterization, HAdV-specific genomic fragments amplified by PCR were purified using the QIAquick PCR Purification Kit (QIAGEN, USA) and subjected to Sanger sequencing at the ACTGene Laboratory (Biotechnology Center, Federal University of Rio Grande do Sul [UFRGS], Porto Alegre, Brazil). The sequences obtained were compared with reference sequences available in the NCBI database. Sequence alignment and phylogenetic analyses were performed using IQ-TREE, MEGA11, and iTOL. Evolutionary relationships were estimated using the composite maximum-likelihood method with 1,000 bootstrap replicates.

## Results

A total of 131 samples were screened for HAdV by PCR. Thirteen (9.9%) tested positive, whereas 118 (90.1%) tested negative. PCR products were visualized by agarose gel electrophoresis, which showed amplicons of 486 bp (Figure 1).

**Figure 1.**
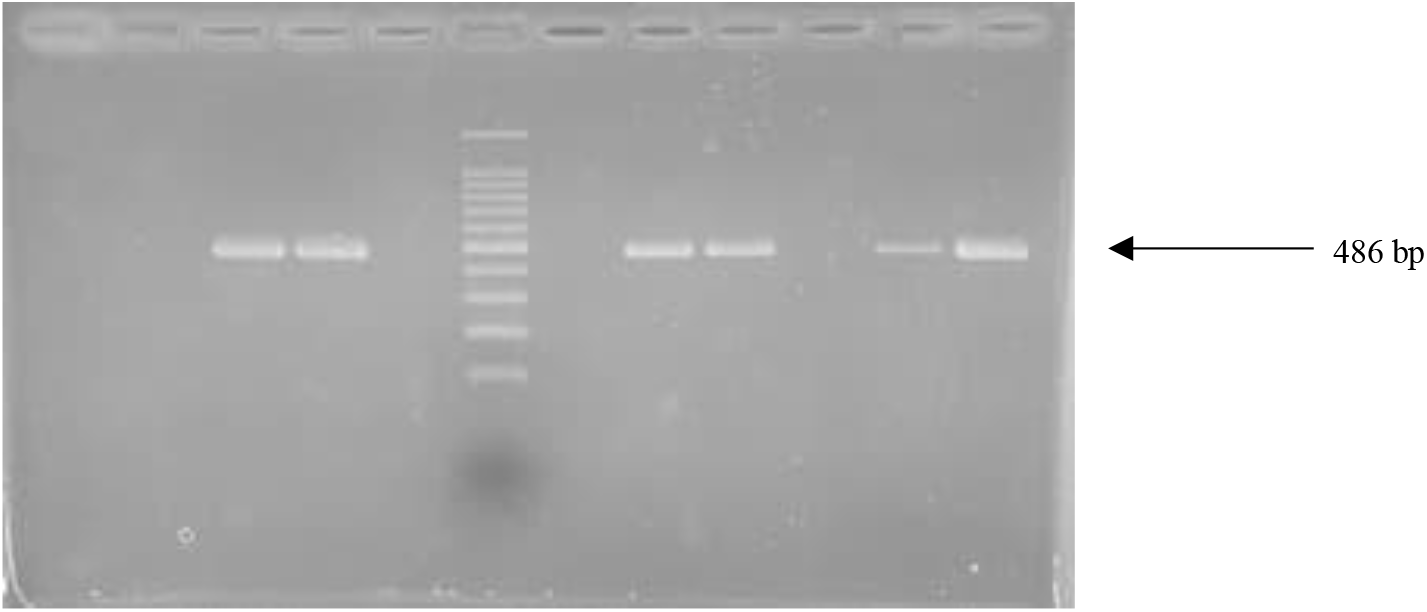
Detection of HAdV by PCR. Lane 1 contains the negative control, and lane 6 contains the 100-bp DNA ladder (Invitrogen, USA). HAdV-positive samples, showing 486-bp fragments, are present in lanes 3, 4, 8, 9, 11, and 12.

Among the 13 HAdV-positive samples, most cases occurred in children aged 0– 3 years. Infants younger than one year accounted for five of the 13 positive cases. Patient ages ranged from 0 to 8 years, as illustrated in Figure 2.

**Figure 2.**
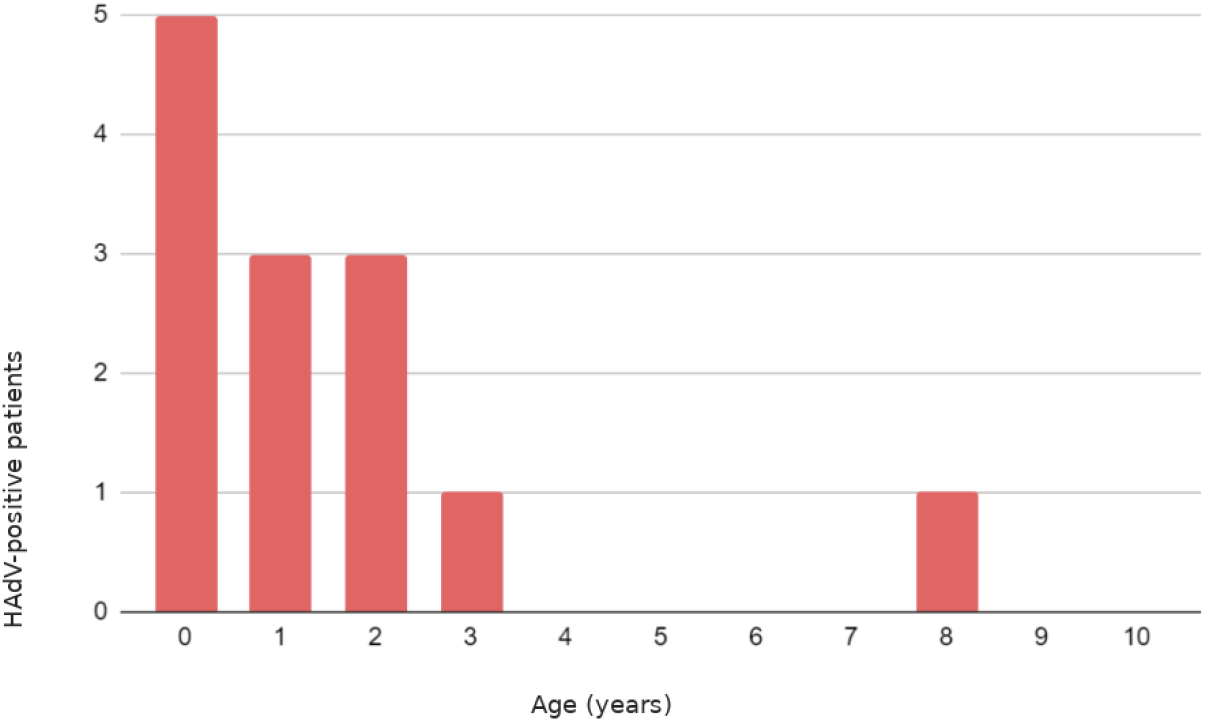
Age distribution of HAdV-positive patients. The chart shows the number of HAdV-positive cases according to patient age. Source: Prepared by the authors, 2024.

Sequence alignment and maximum-likelihood analysis were performed using IQ-TREE. Phylogenetic analysis showed that the Human adenovirus F41 Brazil/2022 strain occupied a distinct position near the base of the phylogenetic tree, indicating genetic differences from the other strains analyzed. This position suggests the possibility of a novel lineage within the F41 group, although the strain showed its closest evolutionary similarity to ON561778.1, Human adenovirus F41 Spain/2022 (Figure 3). Regarding the Human adenovirus B3 Brazil/2022 strains, the analysis showed that Human adenovirus B3 Brazil/2022 1 was evolutionarily close to Asian strains, whereas Human adenovirus B3 Brazil/2022 2 and Human adenovirus B3 Brazil/2022 3 formed a separate cluster, suggesting the possible presence of a distinct or poorly characterized lineage in Brazil (Figure 4).

**Figure 3.**
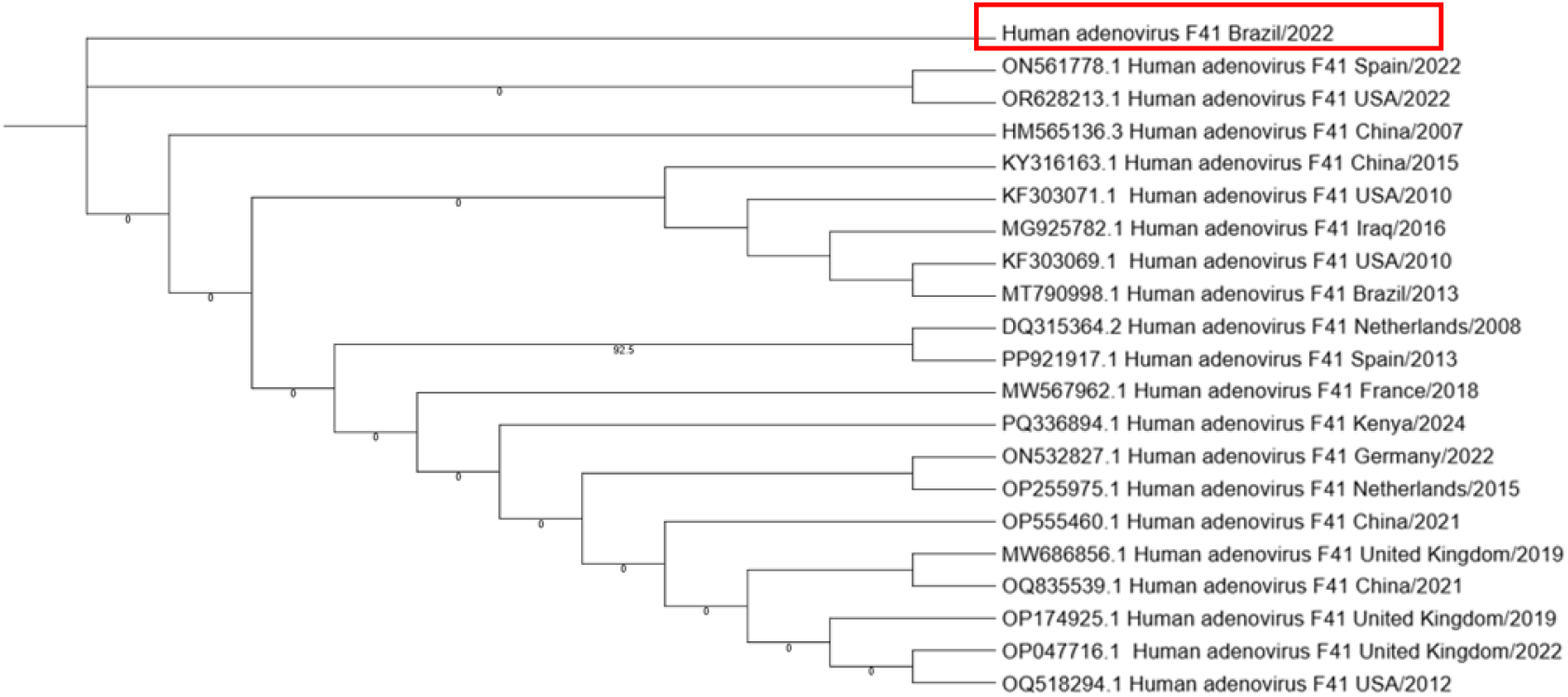
Phylogenetic analysis of the hexon gene of HAdV-F41. The tree was constructed using the composite maximum-likelihood method, with branch lengths expressed as the number of nucleotide substitutions per site and 1,000 bootstrap replicates. The analysis included 21 HAdV-F41 nucleotide sequences retrieved from GenBank (http://www.ncbi.nlm.nih.gov). Evolutionary analyses were performed using IQ-TREE. Source: Prepared by the authors, 2024.

**Figure 4.**
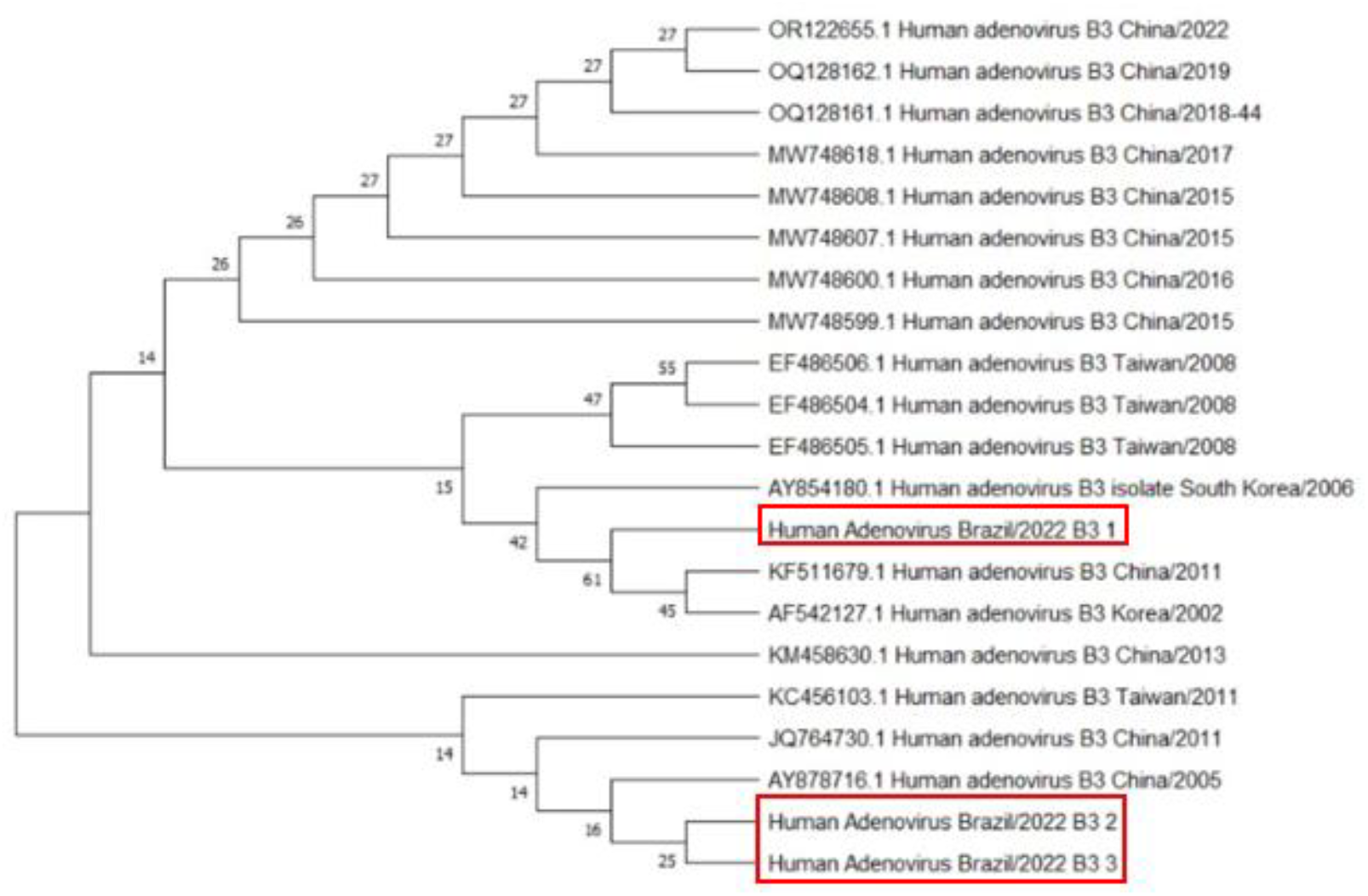
Phylogenetic analysis of the hexon gene of HAdV-B3. The tree was constructed using the composite maximum-likelihood method, with branch lengths expressed as the number of nucleotide substitutions per site and 1,000 bootstrap replicates. The analysis included 21 HAdV-B3 nucleotide sequences retrieved from GenBank (http://www.ncbi.nlm.nih.gov). Evolutionary analyses were performed using IQ-TREE. Source: Prepared by the authors, 2024.

It should be noted that bootstrap values below 70% indicate weak support for the evolutionary relationship represented by a branch in a phylogenetic tree. This means that the same relationship was recovered in fewer than 70% of the bootstrap replicates, suggesting insufficient support for that clade. Branches with support below this threshold should therefore be interpreted cautiously because the corresponding phylogenetic inferences remain uncertain.

## Discussion

Acute gastroenteritis can affect individuals of all ages. In this study, HAdV was investigated as a possible etiological agent of AGE in fecal samples collected in Bahia, and HAdV was detected in the samples analyzed. Previous studies in Salvador reported a prevalence of approximately 2% (Filho et al., 2007), a value considered consistent with the findings of the present study. However, reported frequencies vary considerably, reaching as high as 93.3% in the literature (Tahmasebi et al., 2020). These findings suggest that the virus continues to circulate in the city, highlighting the importance of monitoring its presence during local outbreaks. The authors also emphasize that AGE may not always be clinically diagnosed, potentially allowing these viruses to spread as unrecognized agents.

Analysis of the age distribution of pediatric AGE cases showed that children aged three years or younger accounted for the largest number of cases in this study, including cases potentially associated with HAdV. Previous research in Brazil also reported that children younger than three years accounted for the largest number of deaths from diarrhea and gastroenteritis of presumed infectious origin between 2017 and 2022 (De Melo et al., 2023).

The reasons why younger children appear more frequently among affected patients warrant further investigation. Potential explanations include immune system immaturity (Shaw; Su, 2021), exposure to new pathogens in childcare or school settings, and greater vulnerability to dehydration. These findings are concerning because gastroenteritis can progress to complications such as dehydration (Brandt; de Castro Antunes; da Silva, 2015), making this population particularly vulnerable (Pijnacker et al., 2019). Monitoring viral circulation in pediatric patients is therefore important for the prevention of further clinical complications. The present study also contributes to understanding HAdV epidemiology in Salvador, Bahia, through molecular and cell-culture investigations.

Phylogenetic analyses suggest the possibility of multiple introductions of HAdV-B3 variants into Brazil, with similarities to Asian strains. In addition, the HAdV-F41 Brazil/2022 strain exhibited genetic characteristics that may indicate a unique or poorly studied lineage. However, genetic similarity between Brazilian and Asian sequences does not, by itself, establish independent introduction events or the effects of specific environmental factors. Further studies are therefore needed to better understand the evolution of these strains.

Furthermore, the different geographic origins of the strains may suggest the influence of distinct selective pressures associated with interactions between the virus and human populations, including host genetic variability and regional epidemiological patterns (Gaudin; Goetz, 2021; Gogarten; Townsend, 2005; Sanjuán; Domingo-Calap, 2016). Environmental factors, such as climate and sanitation practices, may also have affected viral behavior and contributed to distinct evolutionary trajectories (Barcellos et al., 2009; Carratalà et al., 2013).

These considerations highlight the complexity of the evolution and dissemination of adenoviruses and other viral pathogens, as well as the need for more detailed studies of the mechanisms involved in the emergence of viral variants. The presence of distinct strains underscores the importance of epidemiological surveillance to better understand the circulation and evolution of HAdV-F41 and HAdV-B3 and to inform strategies for preventing and controlling outbreaks associated with these pathogens. Differences among strains also point to the need for further studies of enteric viruses.

Viral isolation from fecal samples is challenging because of the diversity of microorganisms present in this material, which may interfere with cell culture and complicate identification of the virus of interest (Cao et al., 2022; Eckburg et al., 2005). Despite these difficulties, morphological changes were observed in Vero cells inoculated with HAdV-positive samples, with cytopathic effects appearing four days after inoculation. Detection of viral genetic material by PCR and sequencing, together with the observed cellular changes, provides evidence of HAdV presence in the cultures examined. Nevertheless, confirming productive viral replication requires additional evidence, such as an increase in viral load or infectious titer over the course of culture. Thus, these findings contribute to the characterization of HAdV behavior in Vero cells and provide a basis for further investigation.

The use of Vero cells for HAdV isolation has been described previously; however, the studies consulted did not provide sufficiently detailed descriptions of the cytopathic effects observed to permit direct comparisons (Alarcon-Valdes et al., 2022; Hasler; Wigand, 1978). In the present study, Vero-cell inoculation allowed the observation of morphological changes, contributing to the description of cytopathic effects and the investigation of HAdV biological characteristics (Mello, 2010; Silva et al., 2021).

These findings are relevant to virological research, particularly because they allow comparison with studies using other cell lines and culture conditions. Combining cell culture with molecular methods broadens opportunities to investigate virus–host cell interactions (Zhang; Gauger, 2014), although further analyses are required to establish replication efficiency and potential differences in behavior among the identified variants. Accordingly, the results provide an experimental basis for future investigations into HAdV isolation and characterization (Aoki et al., 2010).

Taken together, the results demonstrate the presence of HAdV in fecal samples from children in Bahia and contribute to the molecular characterization of the variants identified and the investigation of their behavior in cell culture. Genetic relationships between the strains identified here and those reported in other regions highlight the importance of monitoring viral diversity. The observations in Vero cells also provide information relevant to future studies of HAdV isolation and characterization. Nevertheless, further studies are needed to clarify the contribution of these variants to episodes of gastroenteritis and to deepen our understanding of their epidemiological characteristics.

## Data Availability

All data produced in the present work are contained in the manuscript

